# PRESENT SITUATION OF SUICIDE IN BANGLADESH: A REVIEW

**DOI:** 10.1101/2021.02.23.21252279

**Authors:** Most. Zannatul Ferdous, A.S.M. Mahbubul Alam

**Affiliations:** Department of Public Health and Informatics, Jahangirnagar University; Department of Pharmacy, Jahangirnagar University

**Keywords:** Suicides, Bangladesh, methods of suicides, risk factors, suicide in Bangladesh

## Abstract

The most important global cause of mortality is suicide. It is often neglected by researchers, health professionals, health policymakers, and the medical profession. This review was aimed to provide a narrative understanding of the present situation of suicide in Bangladesh based on the existing literature. We conducted a review combining articles and abstracts with full HTML and PDF format. We searched PubMed, PubMed Central, Google Scholar, ScienceDirect and BanglaJOL, google using multiple terms related to suicide without any date boundary and without any basis of types of studies, that is, all types of studies were scrutinized. Finally, 16 articles were selected for review. Report suggested that every day almost 32 people commit suicide in 2019 which was 29 and 30 in 2015 and 2017 respectively. The mortality rate of suicide found 39.6 per 100,000 in Bangladesh. The most common method is hanging followed by poisoning and jumping under the train. The most prevalent age group is age under 40 years. The rate of suicide in children is also increasing. In contrast to most Asian countries, more Bangladeshi women commit suicide than men. The mean age of male and female were 28.86 ± 11.27 years and 25.31 ± 7.70 years respectively. The most common associated factors of suicide are younger age, lower education, students, nuclear family, family history of suicide, use substance, problem in workplace, financial constraints, affair, domestic violence, divorce, and physical illness. Most of the suicidal event occurred at night, followed by morning (6 am–12 am), and evening. It’s a criminal offence in Bangladesh. The source of information is mainly police, forensic reports, media and courts. Till now there is no nationwide survey of suicide is conducted. Besides, suicide surveillance strategy is yet to be established. Suicide is a neglected and under attended public health problem in Bangladesh with few research and paucity of literature. Now nationwide survey conduction and establishment of national suicide surveillance are a time demanded step.

## INTRODUCTION

Suicide is a neglected global public health problem and Bangladesh is not an exception. Unfortunately, suicide all too often fails to be prioritized as a major public health problem. Approximately one million people commit suicide every year worldwide. A 65% increase in the rate of suicide in the past 45 years has been occurred around the world (World Health Organization, 2004). Sixty percent of all cases of suicide in the world occur in Asia and 39.6 per 100,000 in Bangladesh. Suicide can occur at any point in the lifespan, and is the second most frequent, and in some countries the leading cause of death among young people aged 15–24 years (World Health Organization, 2015). In addition, around 20–30 times as many suicide attempts occur (Wasserman, 2001).

World Health Organization (WHO) estimates for the year 2020 and based on current trends approximately 1.53 million people will die from suicide and 10-20 times more people will attempt suicide worldwide which represents on average 1 death per 20 second and 1 attempt every 1-2 seconds (Feroz&et al.,2012; Khan 2005; Ali & et al., 2014). Nowadays Suicide has become a daily occurrence event in Bangladesh. About 10,000 persons are dying by suicide per year in the country (World Health Organization 2014; Mashreky et al., 2013; Shahnaz et al., 2017; Begum et al., 2017a) which was reported by WHO. It is the fourth leading cause of overall injury-related deaths and second important cause of injury-associated death in age groups of 20– 39 years in Bangladesh (Mashreky et al., 2013).

Suicide is a worldwide growing problem crossing culture, geographies, religious, social and economic boundaries, studied by Alexander (Alexander, 2001), Aseltine and DeMartino (Aseltine & Martino, 2004), Johansson et al (Johansson, Lindqvist & Eriksson, 2006). While suicidal behaviour is influenced by several interacting factors - personal, social, psychological, cultural, biological and environmental - depression is the most common psychiatric disorder in people who die by suicide (Cavanagh, Carson, Sharpe & Lawrie, 2003). About half of all individuals in high-income countries who die by suicide have major depressive disorder at the time of their death (Arsenault, Kim & Turecki, 2004; Carroll, Metcalfe & Gunnell, 2016). Moreover, a history of suicide attempts is a robust risk factor for death by suicide (Carroll, Metcalfe & Gunnell, 2016). WHO has estimated that the 26% of the world’s population living in the 11 countries of the WHO South-East Asia Region accounts for 39% of global suicides (World Health Organization, 2017).

So, WHO focuses on suicide prevention and called on the countries to devise national suicide prevention strategies (Arafat, 2017; Caine, 2012, Manton, 2016; Ghanbari, Malakouti, Nojomi, Alavi & Khaleghparast, 2014). Prevention strategies should be based on the risk factors and previous evidences.

Bangladesh is a densely populated country and its economy is rising in South Asia having more incidence rate of suicide than the other Asian countries. Current review reveals that suicide rates in South Asia are high compared to the global average, and still there is a scarce of reliable data on suicide rates in South Asia (Jordans & et al, 2014). Till now there is no national suicide surveillance system. Besides no nationwide study on suicidal risk factors has been yet initiated (Khan 2005; Arafat 2017; Shah et al. 2017; Chowdhury et al. 2018). Furthermore, it is still a criminal offence in the legal system (Arafat, 2017).

Religious and social factors continue to influence the diagnosis and registering of suicides as well as families do not disclose the true nature of the act, for fear of harassment by police and social stigma (Khan, 2005). The literatures on suicide are still limited in Bangladesh context, and there is no comprehensive article on suicide in Bangladesh context. Existing articles focus on the specific method of suicide such as hanging and poisoning as well as the search for the association of mental disorder with suicide is poor. The present narrative review was conducted to provide a comprehensive understanding of suicidal context in Bangladesh based on the existing literature concerning prevalence of suicide, and other suicide metrics like suicidal factors, methods of suicide, impacts of suicide on family and society level, preventive approach, and legal aspects. The review on this topic in Bangladesh can open new prospect to address the national issue to the healthcare professionals, policy makers and planners.

## METHODS AND MATERIALS

We reviewed the literature to get recent information on suicide prevalence, its associated factors and prevention strategies in Bangladesh. Articles related to suicide and Bangladesh was searched for in six electronic databases with searching multiple keywords without any date boundary and without any basis of types of studies, that is, all types of studies were scrutinized. There are few full downloadable articles, and total 30articles were found. After exclusion of repetition, screening, finally selection was done on the basis of inclusion and exclusion criteria and finally 16 articles were selected for review. No meta-analyses were conducted.

### SEARCH STRTEGY

Six electronic databases were systematically searched to retrieve relevant articles. Firstly, we searched PubMed, PubMed Central, Google Scholar, BanglaJOL, ScienceDirect, and google using multiple keyword combinations as follows: [‘Suicide’ or ‘Suicide in male’ or ‘Suicide in female or ‘Suicide in adolescents’ ‘Suicide in children’ or ‘Suicide and its ‘risk factors’] and [‘Suicide and its ‘prevention strategies’ or ‘Suicide and Bangladesh’] and [‘Epidemiology of suicide in Bangladesh’ or ‘Methods of suicide in Bangladesh’ or ‘Prevalence of suicide in Bangladesh’ or ‘Suicide in Bangladesh’]. Databases were searched independently by two reviewers. When there was disagreement (over the identification of relevant studies), studies were reassessed independently and consensus was reached following discussion. Secondly, we applied a snowball method by which we reviewed the bibliographies of all the studies identified as relevant in the preceding step.

### CRITERIA FOR INCLUSION

We included studies that describe suicide in Bangladesh context. The articles were fully downloadable in PDF format and also HTML format were included. Searching was done in PubMed, PubMed Central, Google Scholar, BanglaJOL, ScienceDirect and Google.

### CRITERIA FOR EXCLUSION

Articles other than suicidal topics such as deliberate self-harm and accidental poisoning were excluded. Additionally, for the peer-reviewed articles, we excluded records published in languages other than English, book chapters, conference proceedings, and commentaries. Those studies focus on situation of suicide outside the Bangladesh were also excluded.

### DATA EXTRACTION AND ANALYSIS

Two reviewers selected the included articles. A standard template sheet was developed to capture relevant aspects of the research objective. The data synthesis process involved both narrative and graphic analysis. The process involved reading and re-reading of the included studies by the two reviewers to identify key emerging issues.

## RESULTS

### SERACH OUTCOME

Of 30 articles initially identified from six databases using different keywords, 6 proved irrelevant (not related to situation of suicide in Bangladesh) after the titles were examined and 3 articles were duplicates. Abstracts of the remaining 21 articles were then screened using the inclusion criteria. When it was not clear from the abstract whether a study met them, the full-text article was read. Of these, 5 were excluded (did not meet the inclusion criteria) and 16 articles met the eligibility criteria. Related citations and reference lists of all relevant articles were checked.

### DESCRIPTION OF THE STUDIES

Of the 30 articles 16 met the criteria for inclusion (cross ref: Figure 01). Among the articles, there were 5 original articles, 7 review articles including 4 systematic review and 3 narrative review and 1 thesis as well as 3online newspaper reports.

**Figure 01:**
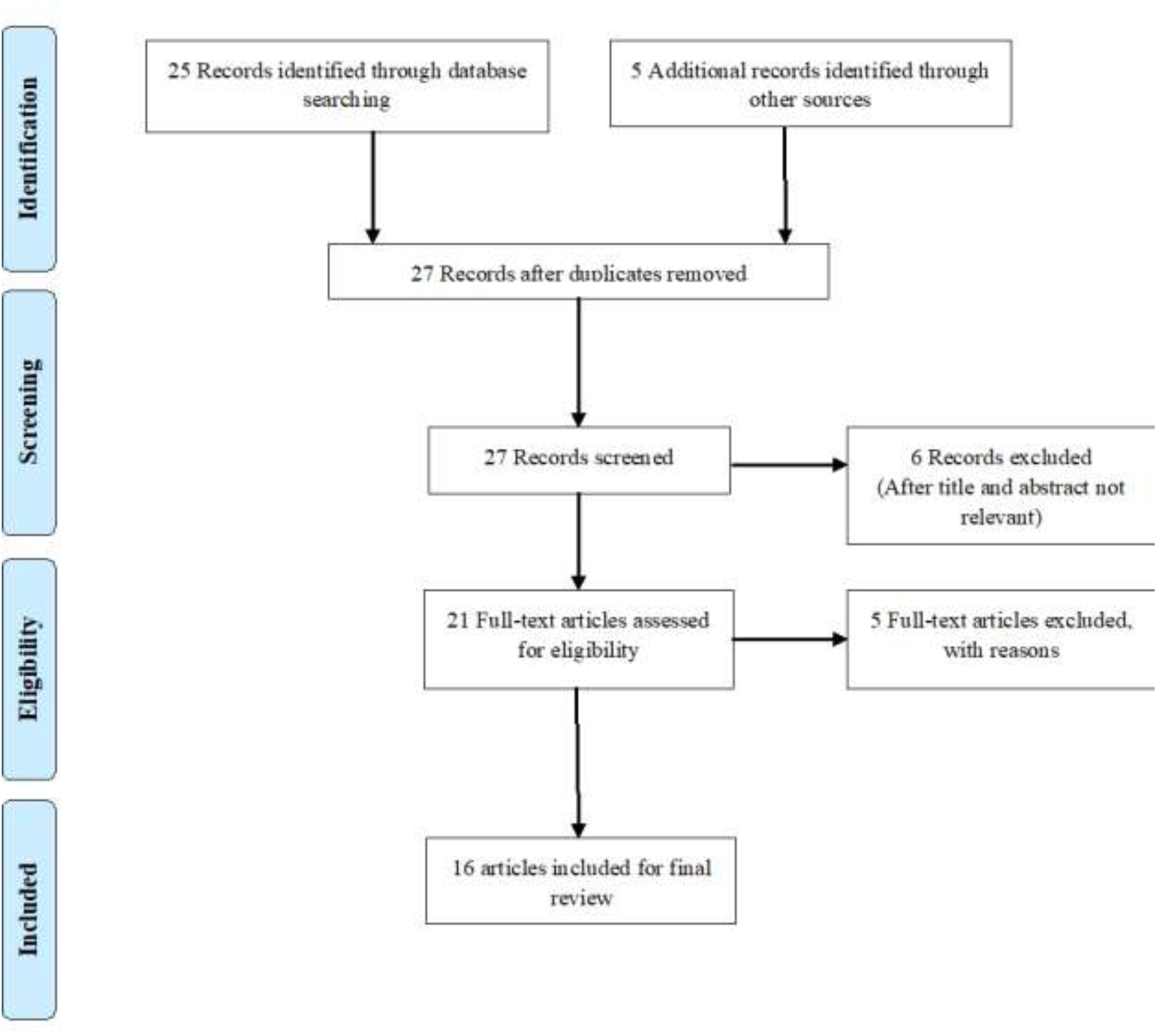
Flow diagram of the study selection process (PRISMA flow diagram)

### PREVALENCE OF SUICIDE

According to the United News of Bangladesh (UNB), in Bangladesh, suicide rates are on the rise for the absence of proper attention to mental health issues and necessary support, and complications in the socialization process (Daily Sun, 2018). In Bangladesh there is no standard reporting system of the suicide prevalence. Besides there is no nationwide survey; and under-reporting is still prevalent in Bangladesh. Like Pakistan, Afghanistan, Nepal, Bangladesh relies mostly on police data which are likely gross underestimations of actual rates. The 2014 report of WHO revealed Bangladesh recorded nearly eight suicides for every 100,000 people (World Health Organization, 2014). The average suicide rate was found 39.6/100,000 population/year in the existing literature (Feroz & et al., 2012; Khan 2005; Ali & et al., 2014; Jordans & et al, 2014). Another report by Shaheed Suhrawardy Medical College Hospital, Dhaka, published in 2010; around 6,500,000 people of Bangladesh are prone to suicide. The rate is 128.08/100,000 populations commit suicide in Bangladesh every year. But this has been noted that the above rate was assumed only based on survey conducted in one union which is not a good reflector of the total suicide rate in the whole country as total suicide rate per year is far less than what was mentioned in the report in the last couple of years (Hasan & Rabby, 2018). The estimated rates of fatal and non-fatal suicidal behavior were 3.29 and 9.86 per 100,000 PYO, respectively.

According to a report by The Daily Star, from 2002 to 2009, 73,389 people committed suicide in Bangladesh. Of these 73,389 people, 31,857 people hanged themselves and 41,532 swallowed poison to commit suicide (Rony, 2018). World Health Organization (WHO) reported, 19,697 people committed suicide in Bangladesh in 2011 (Worldlifeexpectancy.com, 2012). According to Bangladesh Police, which publishes a report annually on suicide incidents, over 11,000 people committed suicide in the country in 2017 which was 9,665 in 2010. In 2016, the total number of suicide incidents in Bangladesh was 10,600 while 10,500 in 2015 and 10,200 in 2014. The prevalence of suicide in Bangladesh shows in figure 02 from the period of 2012 to 2017.

**Figure 02:**
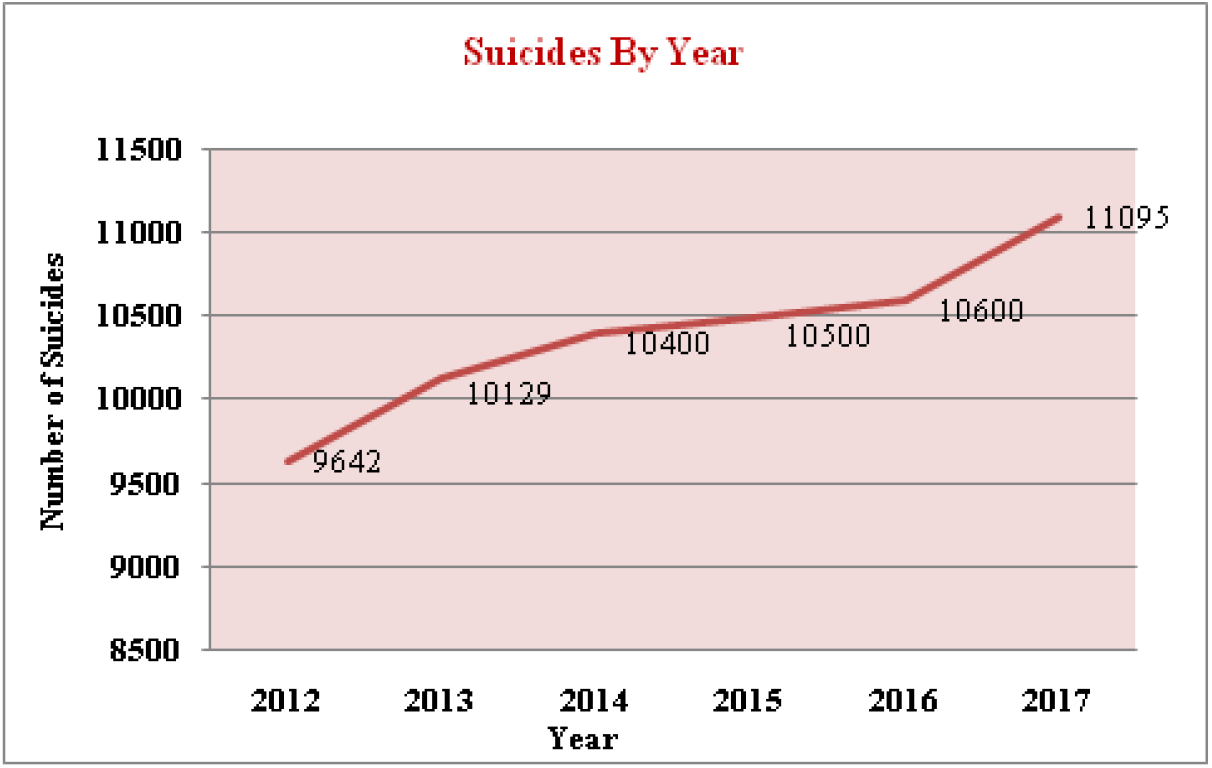
Prevalence of suicides by year (2013-2017); Source: Bangladesh Police.

Police records also show that the number of suicides has increased over the years. The police said, around 6,753 people have committed suicide from January to July this year. This means, every day about 32 people committed suicide which was 29 and 30 in 2015 in 2017 respectively (Rony, 2018; Hasan, 2019). However, the Bangladesh Police could not provide the suicide data for 2018, but stated that 19,094 unnatural deaths occurred in the year, around 11,000 were possibly deaths by suicide and this report was also concurred by several psychiatrists (The Daily Star, 2019). However, the rate of suicide is increasing each year (Hasan & Rabby, 2018). According to psychologists around 11,000 people kill themselves in Bangladesh every year (Hasan, 2019).

### SUICIDE IN PUBLIC AND PRIVATE UNIVERITIES

In last 2 years, suicide rises at public university. According to a report by The Daily Starin 2017, six public university students had died by suicide. Four of them were from Dhaka University (DU) while one each from Jahangirnagar University and Jatiya Kabi Kazi Nazrul Islam University in Mymensingh.

Dhaka University is one of the institutes which have seen an increase in reported suicide cases. While as many as 13 students died by suicide between 2005 and 2016, the same number had died in two years between 2017 and 18. Table 01 shows the number of students who died by suicides in different period of time:

**Table 01:** Rate of suicides in Public University in different time period.

| Years | Number of suicide |
| --- | --- |
| 2005 | 2 |
| 2006 | 2 |
| 2007 | 2 |
| 2012 | 1 |
| 2014 | 1 |
| 2015 | 4 |
| 2016 | 1 |
| 2017 | 4 |
| 2018 | 9 |

**Source: University proctor office**

### AGE AND GENDER DISTRIBUTION

In Bangladesh, a limited number of studies have attempted to quantify suicide rates. Suicides were more prevalent among women than men. Mashreky et al. suggested the rate of suicide was found to be 7.3 per 100,000 population accounting 6.5 in males and 8.2 in females and was highest in the 60+ age group and considerably high among adolescents (Mashreky et al. 2013). According to WHO Global Health Estimates, the suicide rate for 2012 in Bangladesh was 7.8 per 100,000 population with a rate 8.7 in females and 6.8 in males (World Health Organization, 2016). Other study reported causes of female deaths, revealed a growing rate of suicide among adolescent females (Nahar, Arifeen, Jamil & Streatfield, 2015). Reports from national household surveys in 2001 and 2010 and hospital based surveys in 1996–1997 suggested that suicide was the main cause for deaths among adolescent females, accounting for 16–22% of all deaths in this age group (Nahar, Arifeen, Jamil & Streatfield, 2015). Bangladesh Manabadhikar Bastabayan Sangstha, a human rights group of Bangladesh shows that from January 2011 to August 2011, 258 people committed suicide, and of them, 158 were women and the remainder was men. The WHO report also said people of 15-29-year age group are committing suicide most where the rate of suicide is higher among women (World Health Organization, 2018). More worrying is that now even children tend to commit suicide (Daily Sun, 2019)

Salam et al. suggested the risk of suicide was significantly higher by 6.31 times among 15–17 and 4.04 times among 18–24 olds compared to 25–64 years old. Married adolescents were 22 times more likely to commit suicide compared to never-married people (Salam et al., 2017).

However, the age’s ranges from 10 to 78 years (Wu, Chen, & Yip, 2012; Ali & et al., 2014) but most prevalent age group is age under 40 years. The distribution of age was revealed from different study including the most common age is 20-29 years (Feroz & et al., 2012); 20-40 years (Ali & et al., 2014); 20-35 years (Wu, Chen, & Yip, 2012); 21-30 years (Ali & et al., 2014);below 30 years (Talukder, Karim, Chowdhury, Habib, Chowdhury & Perveen, 2014); and 18-40 years (Sarkar, Shaheduzzaman, Hossain, Ahmed, Mohammad & Basher, 2013) age group. The mean age of male was 28.86 ± 11.27 years with age range of 8-70 years and the mean age of female was 25.31 ± 7.70 years with age range of 11-78 years (Ali & et al., 2014).

In contrast to most Asian countries, more Bangladeshi women commit suicide than men (Cross Ref. Table 02). According to the police, around 7,671 women died unnaturally between 2012 and 2017. Among them, 3,444 incidents took place at their parents’ homes while 3,927 incidents occurred at their in-laws’. In contrast, about 9,212 men were victims of unnatural death. However, the Bangladesh Society for the Enforcement of Human Rights (BSEHR) said around 1,374 women committed suicide from 2012 to 2017 whereas 696 men committed suicide in the same period (Hasan & Rabby, 2018). Committing suicide could be divided into three major age groups. First, the teenagers who are more emotional and strongly susceptible to negative inputs are more risk to commit suicide. They were found committing suicide over romantic failures, family disputes, failing exams, or rebuking by parents. For married women, family disputes and insecurity in the workplace was the leading cause. For elderly women, depression and loneliness were the key factors (Rony, 2018).

**Table 02:**
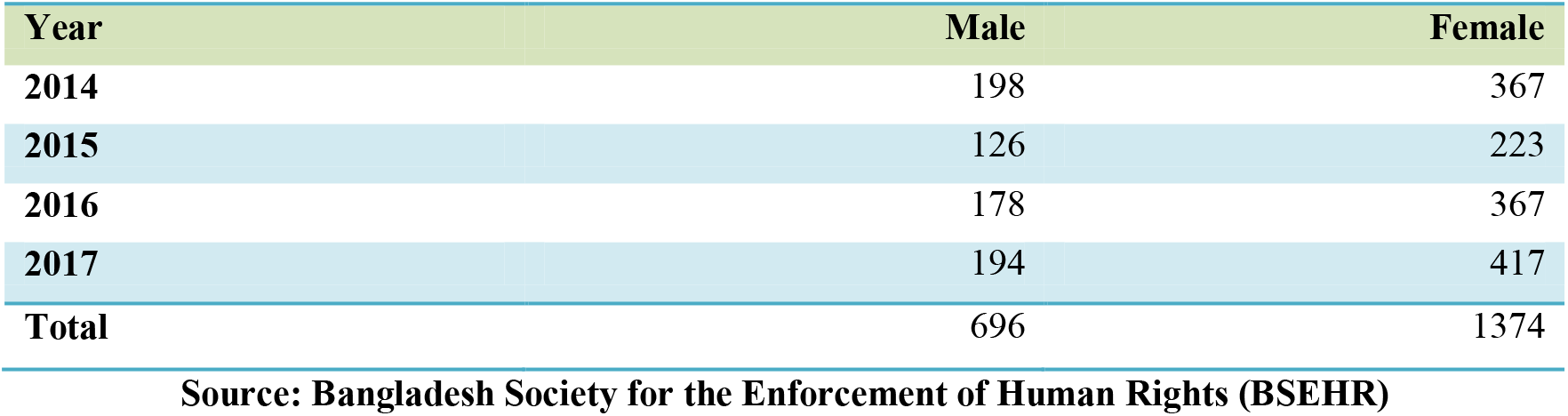
Distribution of suicide in male and female.

In Bangladesh mostly productive age group is being affected by suicide which was revealed by the current review. Oppositely, in worlds’ other parts the older age groups are more prone to suicide. These premature deaths proceeds the economic development of the country and the family structures damage and the end result may prolong to the future generation in regards to physical, psychological, social as well as economic development.

### CHILDREN AT RISK

In Bangladesh, children do not get many opportunities to generate their capacity of thinking and make their own decisions, which, in the long run, results in high dependency on parents. Also, when they become adults, they cannot cope with the complex society and get depressed resulting decision to end life is the solution. In 2019, the numbers appear to be somewhat lower, with 148 deaths of children attributed to suicide, and 10 unsuccessful attempts (Hasan, 2019). Data in figure 03 shows the prevalence of suicides in children.

**Figure 03:**
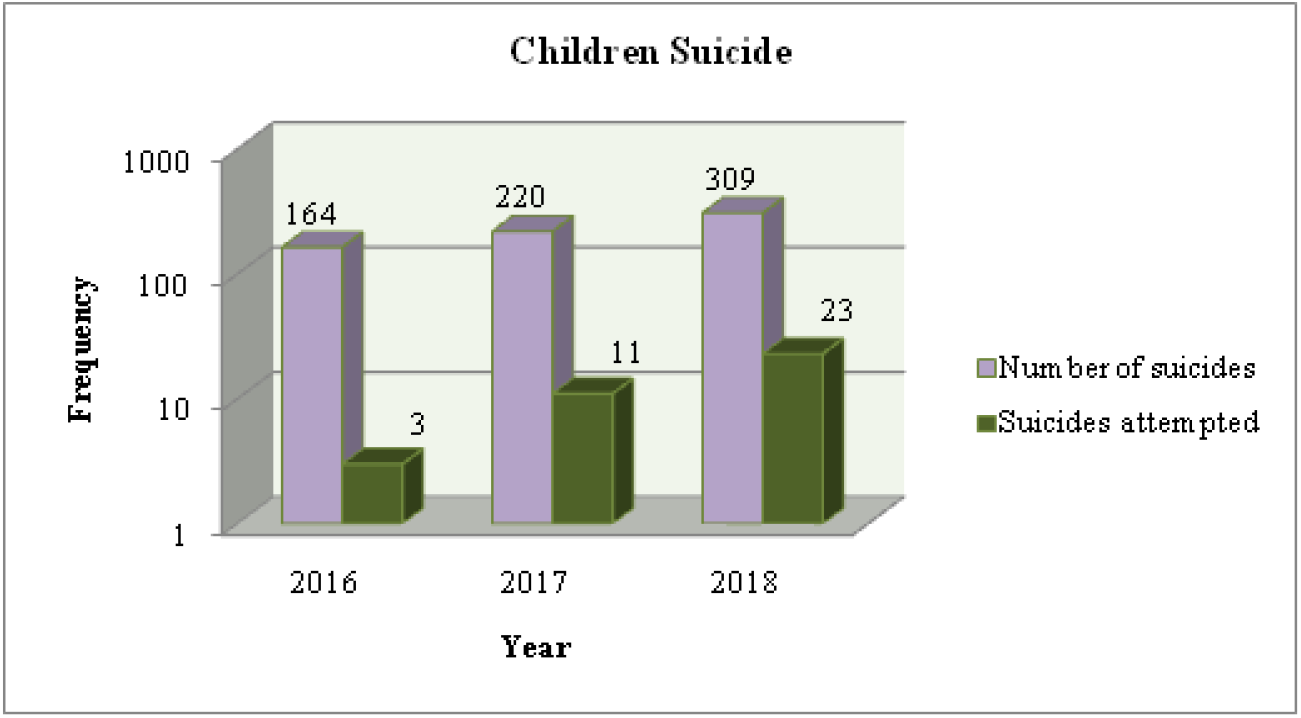
Prevalence of suicides in children, Source: Bangladesh Shishu Adhikar Forum (BSAF)

### PLACE DISTRIBUTION

There were variations in rate in regards to the region of the country. It was found that South-west region of Bangladesh specially Chuadanga, Jenaidah, Kustia and Meherpur districts are the highest (128.8/100000 populations) prevalent area of suicide in Bangladesh (Feroz & et al., 2012; Khan 2005; Ali & et al., 2014). Another survey reported that compared to Chandpur/Comilla district, the risk of suicide was significantly higher in Narshingdi (Salam et al., 2017).

Countries with rates of more than 30/100,000 are considered high rates countries; those with rates between 10 and 29/100,000 as middle rates countries and those with rates <10/100,000 as low rates countries and Bangladesh belongs to the high rates country according to the reported suicidal rate (Khan, 2005; Ali & et al., 2014).

Barisal topped the list in 2017, with a total of 2,585 deaths by suicide. After Barisal, Dhaka district had the highest number of suicides in 2017. That same year, 2,585 people committed suicide in Dhaka, 1,433 people in Rangpur, and 1,197 killed themselves in Sylhet. In 2017, a total of 1,239 suicide cases were reported alone in the Railway Range (areas falling under the jurisdiction of Bangladesh Railways) areas of the country. When compared with districts, the number of suicide in Railway Range area, after Barisal and Dhaka, is more than any other district. Figure 04 shows the distribution of the suicides in the Railway Range area:

**Figure 04:**
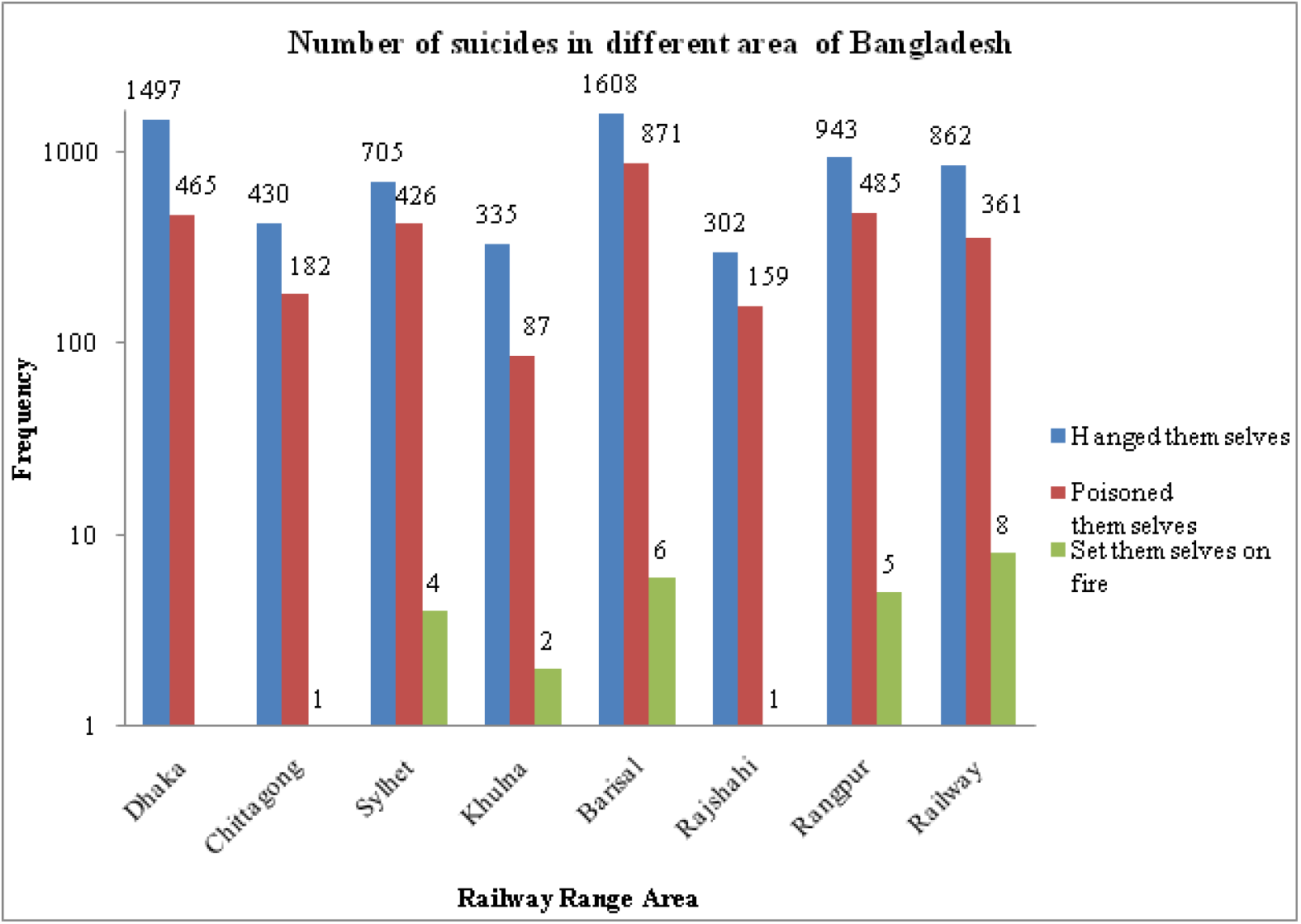
Suicides in different area, Source: Bangladesh Police Headquarters.

### METHODS OF SUICIDE

On the basis of articles methods of suicides vary. The lack of comprehensive articles results paucity regarding existing suicidal methods. Hanging is the most common and preferred method of suicide in Bangladesh (Ahmad & Hossain, 2011). Swallowing poison or ingestion of insecticides is another most common method of suicide in Bangladesh. In urban areas, people follow other methods to commit suicide, such as by an overdose of barbiturate tablets, or by other means. The other mentionable methods are burning, jumping in front of train, fall from the heights, drowning, gun fire, cutting throat (Shah, Ahmed, & Arafat, 2017). Feroz *et al*. found from the community-based survey that the common methods of suicide are hanging and then ingestion of insecticides (Feroz & et al., 2012) while Choudhury *et al*. found that poisoning is the dominant method of suicide (Wu, Chen, & Yip, 2012). There are some other findings that supporting the hanging as the most common method of suicide in Bangladesh (Ali & et al., 2014); then poisoning (Hossain, Rahman & Akhter, 2011). Salam et al. found from census survey that the most common method of suicide was hanging (59%) followed by poisoning (31%), burn (5.1%), drowning (2.6%) and exsanguinations (2.6%) in fatal and these frequencies were reversed for non-fatal suicidal behavior, where poisoning (71.93%) was the most common method followed by hanging (22.81%) (Salam et al., 2017). These methods are consistent with the subcontinent and inconsistent with the west countries (Ali & et al., 2014; Jordans & et al, 2014; Wu, Chen, & Yip, 2012).

However, the choice of method largely depends on their easy availability and accessibility, the high fatality (Feroz & et al., 2012); a traditional culture that influences method choice, the mass media as a powerful cultural channel has played a significant role in the dissemination of suicide methods (Wu, Chen, & Yip, 2012). The numbers of suicides caused by the method firearms are very few in Bangladesh (Arafat, 2017).

According to data from Police Headquarters, among the 11,095 people who committed suicide in 2017, 569 of them hanged themselves, 3,467 took poison, and 59 people set themselves on fire (Rony, 2018).

In Bangladesh majority of people live in villages. Still our economy is agricultural-based. Both hanging and poisoning are preferable to existing culture as well and insecticides are lethal, easily available, accessible, poor storage can be the causes for choosing (Feroz & et al., 2012, Ali & et al., 2014; Wu, Chen, & Yip, 2012; Talukder, Karim, Chowdhury, Habib, Chowdhury & Perveen, 2014). Regarding hanging Dopatta (orna) was the most common ligature material (Wu, Chen, & Yip, 2012; Talukder, Karim, Chowdhury, Habib, Chowdhury & Perveen, 2014); wherein report from another study rope was the most common ligature material in hanging (Hossain, Rahman & Akhter, 2011).

### FACTORS ASSOCIATED WITH SUICIDE

World Health Organization has reported stress, mental illness, unemployment and substance abuse (World health organization, 2014) are the major factors for suicide. A cross-sectional study conducted in Bangladesh reported patients who were younger age, females, lower education, students, nuclear family, family suicide history, use substance showed trend to have higher suicidal ideation (Mali, Akter & Arafat, 2018). Depression, lack of capability to cope up with a situation, absence of proper support and care, complications in socialization process and negligence to mental health issues are responsible for the suicidal tendency (Daily Sun, 2019). Other common causes reported by different newspaper like anxiety, significant academic pressure, family problems, trauma and societal pressure (Rony, 2018). A woman is sexually harassed by a man, the victim feels ashamed and sometimes makes a suicide attempt because she thinks her dignity has completely been ruined (Rony, 2019). In addition, other risk factors include problem in workplace, financial constraints, affair, domestic violence, divorce, physical illness (Shah, Ahmed, & Arafat, 2017).

### TIME OF SUICIDE

The time for taking the fatal suicidal behavior is not fixed. A six-month paper content analysis in Bangladesh reported that most of the suicidal event occurred at night, followed by morning (6 am–12 am), evening, noon (12 pm–4 pm) and afternoon (4 pm–6 pm), night (6 pm–12 am) and late night (12 am–6 am)(Shah, Ahmed, & Arafat, 2017).

### REASON BEHIND SUICIDES ON SHARP RISE

According to experts, in general the reason behind most of the suicide attempts are those issues ranging from depressive disorders, bipolar disorders, anxiety disorders, alcohol and other substance abuse, schizophrenia. Besides others like psychoses, personality disorders, aggression, impulsivity, and hostility, hopelessness, heredity, childhood trauma, past attempts, and ideation. Bangladesh Society for the Enforcement of Human Rights (BSEHR) Executive Director said impulsivity – hopelessness among youths, drug abuse, childhood trauma and past attempts and ideation are the leading causes of suicide attempts. In addition, man lead very busy lives, they do not give time to our friends and family. People tend to feel isolated that way. All of this is increasing the rate of suicide in the country (Hasan & Rabby, 2018).

### SUICIDE AND SDGs

Sustainable Development Goal (SDG) 3 aspires to ensure health and well-being for all at all ages. One aspect of this goal is mental diseases. In case of mental health addressing suicide is more significant. SDG goal 3 indicator 4 (3.4) addresses suicide. As like: Indicator 3.4.2 Suicide mortality rate (per 100,000 populations). Suicide is the most common cause of unnatural death in Bangladesh with higher proportion of women having tendency to commit suicide. While mental disorders in the form of depression and anxiety are common causes of suicide in many societies. There are some proximate causes of suicide for women in Bangladesh such as physical and domestic violence. Suicide mortality rate has been declining from 7 per 100,000 populations in 2000 to 5.5 in 2015. However data reported by Public Security Division (PSD) shows the rate to be 7.1 in 2015 (General Economic Division, 2018).

### WARNING SIGNS OF SUICIDE

World Health Organization reported some of the warning signs of suicide are: threatening to kill oneself; making indirect statements such as “no one will miss me when I am gone”, or referring to death as a place to go; looking for ways to kill oneself (e.g. seeking access to pills, firearms, pesticides); describing suicide as a solution to a problem; giving away valued possessions; saying goodbye to close friends or family members (World Health Organization, 2014).

### LEGAL ASPECTS

Suicide is considered as the criminal offence in Bangladesh that is also prevailed in some other countries also but in many developed countries it is not considered as criminal offence (Khan, 2005,). Religious factors and social factors as well as legal consequences hinder the suicide disclosures.

### SUPPORTIVE ORGANIZATIONS

There are no government institutions which support people who struggle with issues which may lead to suicide. There are no help lines. The only assistance comes from private NGOs like ‘Kaan Pete Roi’. This is the country’s first emotional support centre helpline. Kaan Pete Roi has listed 22 facilities in Dhaka city where people can find support, including educational institutions and clinics (Hasan & Rabby, 2018).

### REPORTING SYSTEM AND ORIGINAL SOURCE OF INFORMATION

Although the reported suicide rate in Bangladesh is high compared to the global average, there is a paucity of reliable data as there is no suicide surveillance and even nationwide survey has yet to be done (Khan, 2005; Wu, Chen, & Yip, 2012). The source of information are based on police, forensic medicine data, media, court, and few other institutional reports such as those from hospitals as well as the focused study on selected population (Feroz & et al., Khan, 2005; 2012, Ali & et al., 2014).

### IMPACTS OF SUICIDE

Suicide impacts on the world’s most vulnerable populations and is highly prevalent in already marginalized and discriminated groups of society. It is not just a serious public health problem in developed countries but most suicides occur in low- and middle-income countries where resources and service are often scarce and limited for early identification, treatment and support of people in need (Daily Sun, 2019). These striking facts and the lack of implemented timely interventions make suicide a global public health problem that needs to be tackled imperatively (Daily Sun, 2019).

The suicide may impact on society like social disorder, extreme poverty, frightening and alarming situation, decrease social status, upward mobility, defame of the society and anarchy in society. It may also impacts on family like family breaking down, mental conflict, financial crisis, family lose intimacy, depression in family members, tendency to commit suicide and traumatic stress and grief (Ara, Uddin & Kabir, 2016).

### PREVENTIVE APPROACH

Incidents of suicide are increasing worldwide. According to WHO, across the globe, more people die from suicide than in warfare (Hasan & Rabby, 2018). WHO published a report titled “Preventing suicide: A global imperative” where it noted that suicide rates worldwide had fallen Bangladesh ranked 10th on the list; with nearly eight suicides for every 100,000 people (World Health Organization, 2014). So, this scenery gives the idea what we have to do at present. Suicide is a preventive public health problem. Till now support from Govt. is relatively low. A specialized clinic “Suicide Prevention Clinic (SPC)” has been started at department of Psychiatry of Bangabandhu Sheikh Mujib Medical University in September 2016 to provide the specialized care of the patients with suicidal behavior (Arafat, 2017). Society for Suicide Prevention Bangladesh (SSPB)was formulated in 2016 with the hope to start the prevention activities in the country freshly. ‘Kaan Pete Roy’is hot line-based NGOs suicide prevention activity which is also known as Bangladesh–Be frienders, is an initiative of International Association for Suicide Prevention (IASP) (Arafat, 2017). It is the only active platform to help people in need (Hasan & Rabby, 2018).

WHO Bangladesh is working to develop mental health, and suicide prevention material as part of a training package for school staff (Hasan & Rabby, 2018). The WHO, the NIMH (National Institute of Mental Health and Hospital), the Directorate General of Health Services, and various stakeholders are also collaborating to train primary healthcare service providers, based on WHO mhGAP (Mental Health Gap Action Programme) (World Health Organization, 2014).

### FUTURE DIRECTIONS AND RECOMMENDATIONS

We should address mental health issues more seriously. It is suggested that inclusion of subject on mental health in school curricula is more in need. Further, employing adequate psychologists across the country is necessary. In addition, creating awareness among mass people about mental health is also necessary. The government urges to take comprehensive plan to eliminate the reasons behind suicide in Bangladesh as well as to identify suicide as public health issue.

Other activities include establishing dedicated government unit to study, and monitor suicide trends, set up suicide hotlines, address mental illness more vigorously, schools and healthcare workers trained to identify signs, and help cope and provide follow-up care to those who attempted to commit suicide (Hasan & Rabby, 2018).

## CONCLUSION

Bangladesh is a densely populated middle income country with high suicide rates. Existing literatures reveal the rate of suicide is increasing than before. In our country suicide is an under attended public health problem. The research on suicide is few and there is paucity of literature. Establishment of national suicide surveillance is now a time demanded step which assess the need scientifically. At the end responsible authority should take necessary steps to address it.

## Data Availability

Not Applicable

